# Opioid Prescribing Mediating State Policy Intervention Effects on Drug Overdose Mortality

**DOI:** 10.1101/2021.05.13.21257109

**Authors:** Jacob James Rich, Robert Capodilupo

## Abstract

The Centers for Disease Control and Prevention reported 70 630 drug overdose deaths for 2019 in the United States, 70.5% of which were opioid-related. Preliminary estimates now warn that drug overdose deaths likely surpassed 86 000 during 2020. Despite a 57.4% decrease in opioid prescribing since a peak in 2012, the opioid death rate has increased 105.8% through 2019, as the share of those deaths involving fentanyl increased from 16.4% to 72.9%. This letter seeks to determine whether the opioid prescribing and mortality paradox is robust to accepted methods of causal policy analysis and if prescribing rates mediate the effects of policy interventions on overdose deaths. Using log_e_-log_e_ ordinary least squares with three different specifications as sensitivity analyses for all 50 states and Washington, DC for the period 2001-2019, the elasticities from the regressions with all control variables report operational prescription drug monitoring programs (PDMPs) reduce prescribing rates 8.7%, while mandatory PDMPs increase death rates from opioids 16.6%, heroin and fentanyl 19.0%, cocaine 17.3% and all drugs 10.5%. There is also weak evidence that recreational marijuana laws reduce prescribing, increases in prescribing increase pain reliever deaths, pill mill laws increase cocaine deaths, and medical marijuana laws increase total overdose deaths, with demographic variables suggesting states with more male, less non-Hispanic white, and older citizens experience more overdoses. Weak mediation effects were observed for pain reliever, cocaine, and illicit opioid deaths, while broad reductions in prescribing have failed to reduce opioid overdoses.

---

The Centers for Disease Control and Prevention reported an unprecedented 70 630 drug overdose deaths for 2019 in the United States, 70.5% of which were opioid-related. Preliminary estimates now warn that drug overdose deaths likely surpassed 86 000 during 2020. Although COVID-19 likely played a role, this development has followed a decades-long trend that has motivated government action. Despite a 57.4% decrease in opioid prescriptions from pharmacies since a peak in 2012, the opioid death rate has increased 105.8% through 2019, as the share of those deaths involving fentanyl increased from 16.4% to 72.9%.^[1]^ Recent systematic, difference-in-differences research of state-level policy interventions suspects “The opioid paradox may arise from the success—not failure—of state interventions to control opioid prescriptions.”^[1]^ However, the authors did not estimate whether mortality predictively followed reductions in prescribing. This letter seeks to determine whether the opioid prescribing and mortality paradox is robust to other accepted methods of causal analysis,^[2][3][4]^ which conveniently produce mediation evidence to determine whether the policies affected various types of drug-related mortality through their effects on opioid prescribing.^[5]^

## Methods

In harmony with previous pooled cross-sectional studies estimating the effects of multiple policy interventions simultaneously,^[2][3][4]^ we first use log_e_-log_e_ ordinary least squares to estimate the effects nine policy indicator “dummy” variables^[1][2]^ had on various drug overdose mortality rates for all 50 states and Washington, DC for the period 2001-2019. We then use the same process to determine whether the policies had an effect on per capita milligram morphine equivalent (MME) “prescribing rates” from pharmaceutical and narcotic treatment retailers calculated from Drug Enforcement Administration reports. Finally, we regress mortality on the policies and prescribing simultaneously. For the independent variables that are significant in the first two models, we determine whether their coefficients reduce in magnitude or become less significant after introducing prescribing in the third model, which signals a mediation effect.^[5]^ Our policy dates match the JAMA Network literature,^[1][2][3][4]^ except for correcting the Delaware operational PDMP date to March 2012.

To reduce the chance of spurious results, we conduct sensitivity analyses with three different specifications containing various combinations of control variables.^[2][3][4]^ The first controls for unobserved factors with time and fixed effects; the second adds demographic variables from similar studies;^[1][2][3][5]^ and the third adds linear time trends. Every regression clusters robust standard errors around the state variable and weights for population.^[2][3][4]^ Estimates in the form of elasticities are reported from the third specification with asterisks for the least-significant specification *P*-value.

## Results

Figure 1 shows the average state-level death rates from regulated and illicit opioids like heroin and fentanyl before and after operational prescription drug monitoring program (PDMP) adoption. Table 1 reports operational PDMPs reduce prescribing rates 8.7%, while mandatory PDMPs increase death rates from all opioids 16.6%, illicit opioids 19.0%, cocaine 17.3%, and all drugs 10.5%. If significance requirements are relaxed to *P* < 0.1, recreational marijuana laws reduce prescribing, more prescribing increases pain reliever deaths, pill mill laws increase cocaine deaths, medical marijuana laws increase total overdose deaths,^[2]^ and states with more male, less non-Hispanic white, and older citizens experience more overdoses.

**Table 1.** Regression Results for Models 1, 2, and 3.

|  | Prescribing Rate (MMEs) | Opioid Death Rate (T40.0-40.4 & T40.6) |  | Pain Reliever Death Rate (T40.2) |  | Illicit Opioid Death Rate (T40.1 & T40.4) |  | Cocaine Death Rate (T40.6) |  | Drug Overdose Death Rate (X40-44, X85, & Y40-44) |  |
| --- | --- | --- | --- | --- | --- | --- | --- | --- | --- | --- | --- |
| Model | 1 | 2 | 3 | 2 | 3 | 2 | 3 | 2 | 3 | 2 | 3 |
| Prescribing Rate (MMEs) |  |  | 0.049 |  | 0.470 ° |  | -0.166 |  | -0.260 |  | 0.130 |
| PDMP Mandatory Law | -0.041 | 0.164 * | 0.166 * | 0.039 | 0.058 | 0.197 * | 0.190 * | 0.183 * | 0.173 * | 0.100 * | 0.105 * |
| PDMP Operational Law | -0.087 * | 0.010 | 0.014 | -0.096 | -0.055 | 0.037 | 0.022 | -0.000 | -0.023 | 0.002 | 0.013 |
| Pill Mill Law | -0.001 | -0.039 | -0.039 | -0.086 | -0.086 | 0.052 | 0.052 | 0.187 | 0.187 ° | -0.027 | -0.027 |
| Day Restrictions Law | 0.008 | 0.095 | 0.094 | -0.117 | -0.121 | -0.011 | -0.010 | 0.023 | 0.025 | 0.067 | 0.066 |
| Good Samaritan Law | -0.070 | 0.001 | 0.004 | -0.004 | 0.028 | 0.121 | 0.110 | 0.073 | 0.055 | -0.002 | 0.006 |
| Naloxone Access Law | 0.084 | 0.099 | 0.095 | 0.159 | 0.119 | 0.186 | 0.200 | 0.058 | 0.080 | 0.051 | 0.040 |
| Medicaid Expansion Law | -0.011 | 0.081 | 0.082 | 0.083 | 0.088 | -0.082 | -0.084 | 0.030 | 0.028 | 0.047 | 0.049 |
| Medical Marijuana Law | 0.016 | 0.080 | 0.079 | 0.005 | -0.002 | 0.026 | 0.029 | 0.058 | 0.062 | 0.099 ° | 0.097 ° |
| Recreational Marijuana Law | -0.059 ° | -0.030 | -0.027 | 0.039 | 0.067 | 0.131 | 0.121 | 0.004 | -0.011 | -0.026 | -0.018 |
| Percent Male<sup>‡</sup> | -5.177 | 23.36 | 23.62 | 12.49 | 14.93 | 34.10 | 33.25 | 30.81 ° | 29.53 ° | 21.66 * | 22.33 * |
| Percent non-Hispanic White<sup>‡</sup> | 1.975 | -8.545 ° | -8.644 ° | -4.225 | -5.154 | -6.647 | -6.311 | -3.422 | -2.898 | -7.632 * | -7.889 |
| Average Age<sup>‡</sup> | -4.023 | 20.19 * | 20.39 ° | 17.57 | 19.46 | 28.95 | 28.29 | 19.42 ° | 18.41 ° | 14.50 | 15.03 |
| Percent in Poverty<sup>‡</sup> | -0.004 | -0.076 | -0.076 | 0.018 | 0.020 | -0.022 | -0.023 | -0.167 | -0.168 | -0.029 | -0.028 |
| Median Income (2019 Dollars)<sup>‡</sup> | 0.146 | -0.362 | -0.369 | -0.176 | -0.245 | -0.816 | -0.792 | -0.410 | -0.371 | -0.203 | -0.222 |
| Medical Insurance Rate<sup>‡</sup> | 0.142 | -1.765 | -1.772 | -0.464 | -0.531 | 0.598 | 0.621 | -1.263 | -1.228 | -1.366 | -1.385 |
| n | 969 | 969 | 969 | 969 | 969 | 965 | 965 | 943 | 943 | 969 | 969 |
| Adjusted R<sup>2</sup> | 0.984 | 0.901 | 0.901 | 0.88 | 0.883 | 0.917 | 0.917 | 0.914 | 0.915 | 0.908 | 0.908 |

All results are from Specification 3 with \*, \*\*, \*\*\*, and ° marking significance at the $P < 0.05$ , $P < 0.01$ , $P < 0.001$ , and $P < 0.1$ levels for all specifications, respectively, with similar estimates and confidence intervals that do not include zero.
<sup>‡</sup> only included in Specifications 2 and 3.

**Figure 1.**
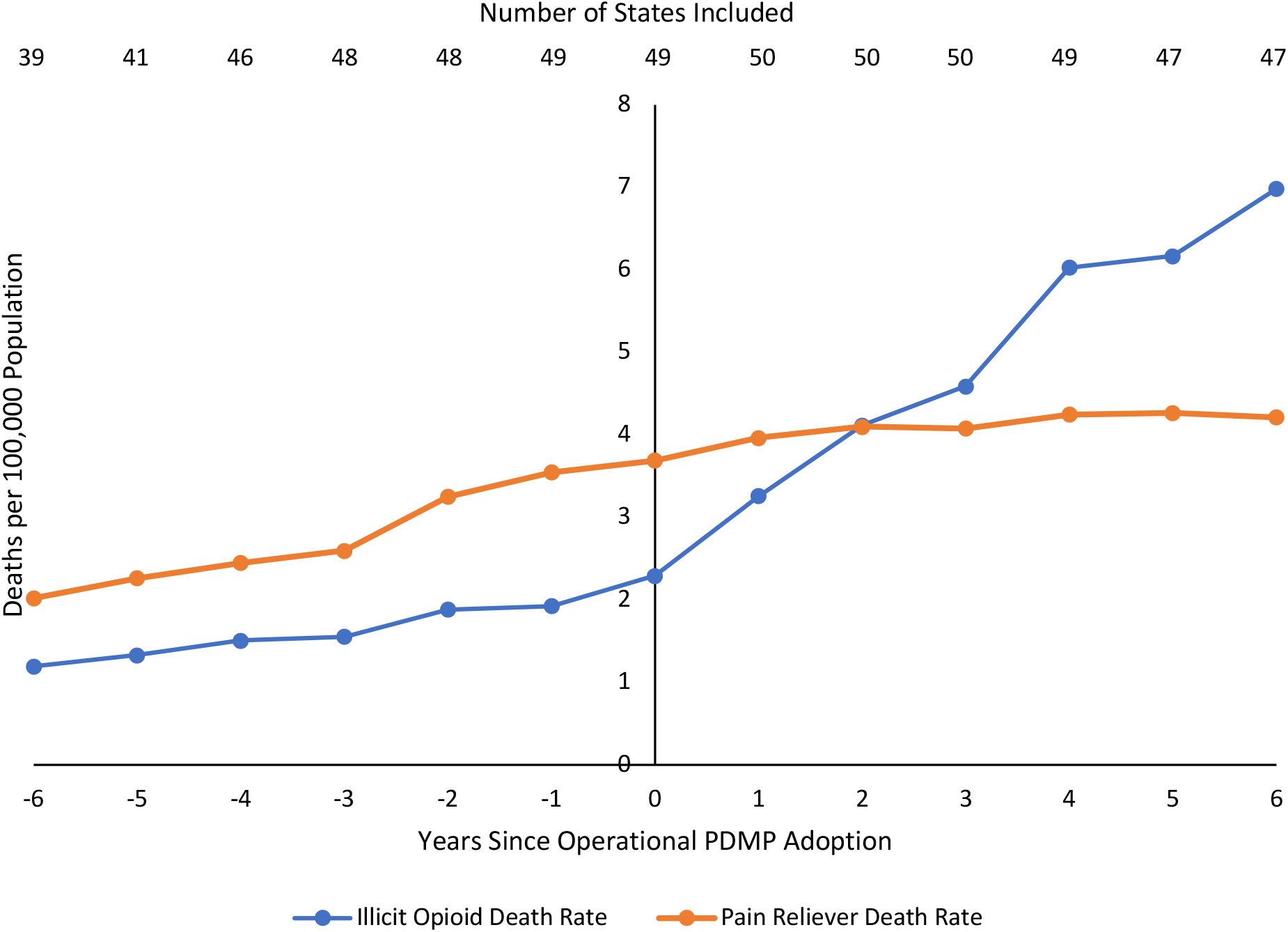
Regulated and Illicit Opioid Overdose Death Rates Since Operational PDMP Adoption.

## Discussion

Opioid policy interventions have successfully achieved their intermediate goal of reducing prescribing, which possibly mediated reductions in pain reliever overdoses, but compromised the ultimate goal of reducing total drug overdose mortality by weakly mediating more poisonings from illicit drugs. With prior research suspecting that the proliferation of drug enforcement leads to more concentrated substances in black markets,^[6]^ the 244.9% increase in the cocaine death rate since 2012 was likely due to interventions increasing the proportion of those deaths involving fentanyl from 4.1% to 63.8%. Regardless, broad reductions in prescribing have failed to reduce opioid-related deaths.

## Data Availability

All data are publicly available: total drug poisoning mortality with subsets for opioid, pain reliever, heroin and fentanyl, and cocaine (X40-44, X60-64, X85, or Y10-14; subsets respectively T40.0-40.4 and 40.6, T40.2, T40.1 and T40.4, T40.5), age, sex, and race population estimates are sourced from the Centers for Disease Control and Prevention (CDC) Wide-ranging Online Data for Epidemiologic Research (WONDER) NVSS database; opioid policy dates from Lee et al. (2021); marijuana policy dates from Anderson et al. (2019); health insurance coverage, median income, and poverty from the US Census Bureau.

https://wonder.cdc.gov/controller/saved/D77/D76F559

https://jamanetwork.com/journals/jamapediatrics/fullarticle/2737637

https://www.census.gov/data/tables/time-series/demo/income-poverty/historical-income-households.html

https://www.census.gov/data/tables/time-series/demo/health-insurance/historical-series/hic.html

